# Waning of the Humoral Response to SARS-CoV-2 in Pregnancy is Variant-Dependent

**DOI:** 10.1101/2021.11.03.21265478

**Authors:** Romina Plitman Mayo, Tal Raz, Bar Ben David, Gila Meir, Haim Barr, Leonardo J. Solmesky, Rony Chen, Ana Idelson, Lucilla Zorzetti, Rinat Gabbay-Benziv, Yuval Jaffe Moshkovich, Tal Biron-Shental, Gil Shechter-Maor, Hen Yitzhak Sela, Itamar Glick, Hedi Benyamini Raischer, Raed Salim, Yariv Yogev, Ofer Beharier, Debra Goldman-Wohl, Ariel Many, Michal Kovo, Simcha Yagel, Michal Neeman

## Abstract

**Importance:** **The** SARS-CoV-2 alpha variant posed increased risk for COVID-19 complications in pregnant women. However, its impact on the maternal humoral response and placental IgG transport remains unclear.

**Objective:** To characterize the maternal humoral waning and neonate immunity acquired during the 3^rd^ COVID-19 wave in Israel, dominated by the Alpha variant, as compared to earlier Wildtype infections and humoral response to vaccination across gestation.

**Design:** Maternal and fetal blood serum were collected at delivery since April 2020 from parturients. Sera IgG and IgM titers were measured using the Milliplex MAP SARS-CoV-2 Antigen Panel supplemented with additional HA-coupled microspheres.

**Setting:** A nationwide multicenter cohort study on SARS-CoV-2 infections and vaccination during pregnancy.

**Participants:** Expectant women presenting for delivery were recruited at 8 medical centers across Israel and assigned to 3 primary groups: SARS-CoV-2 positive (*n* = 157) and fully vaccinated during pregnancy (*n* = 125), and unvaccinated noninfected controls matched to the infected group by BMI, maternal age, comorbidities and gestational age (*n =* 212). Eligibility criteria included pregnant women without active COVID-19 disease, age ≥18 years and willingness to provide informed consent.

**Main Outcome(s) and Measure(s):** Pregnant women’s humoral response is dependent on the SARS-CoV-2 strain.

**Results:** The humoral response to infection as detected at birth, showed a gradual and significant decline as the interval between infection/vaccination and delivery increased. Significantly faster decay of antibody titers was found for infections occurring during the 3^rd^ wave compared to earlier infections/vaccination. Cord blood IgG antigens levels correlated with maternal IgG. However, cord IgG-HA variance significantly differed in SARS-CoV2 infections as compared to the other groups. No sexual dimorphism in IgG transfer was observed. Lastly, high fetal IgM response to SARS-CoV-2 was detected in 17 neonates, all showing elevated IgM to N suggesting exposure to SARS-Cov-2 antigens.

**Conclusions and Relevance:** Infections occurring during the 3^rd^ wave induced a faster decline in humoral response when compared to Wildtype infections or mRNA BNT162b2 vaccination during pregnancy, consistent with a shift in disease etiology and severity induced by the Alpha variant. Vaccination policies in previously infected pregnant women should consider the timing of exposure along pregnancy as well as the risk of infection to specific variants of concern.

**Key Points:** *Question:* What is the difference in the maternal-fetal humoral response between Alpha variant and SARS-CoV-2 Wildtype infections?

*Findings:* In this nationwide multicenter study including 494 pregnant women, the maternal humoral response to Alpha variant infection was weaker and shorter when compared to Wildtype infections. Placental transport compensated for the maternal waning of immunity. Fetal sex did not affect humoral response.

*Meaning:* Vaccination policies should be adjusted to account for the timing of infection and the SARS-CoV-2 variant.

## Introduction

SARS-CoV-2 variants of concern (VOC) associated with increased transmissibility, severity or possible immune evasion have rapidly dominated COVID-19 surges around the world ^1^. Massive whole-genome SARS-CoV-2 sequencing as well as S gene target failures in PCR testing revealed the extremely rapid expansion of the alpha (B.1.1.7) lineage in the UK, consistent with its higher transmissibility, a 50% to 100% higher reproduction number, increased disease severity and increased mortality ^2^. The Alpha variant has accumulated 23 mutations, of which 14 are non-synonymous and six synonymous ^3-5^, involving changes in the S protein, in the receptor-binding domain (RBD), and in the nucleocapsid (N) ^4^. Of these changes 47% occurred in the gene encoding for the S protein, leading to alterations in the interaction with the human angiotensin-converting enzyme-2 (hACE2) receptor, thereby increasing the infection rate ^3^. Consequently, the Alpha variant was the first to be designated as a VOC (2020/12/01) by Public Health England in the fall of 2020 ^6^.

In Israel, the 3^rd^ COVID-19 wave (2021/01/01 – 2021/04/01) was dominated by the Alpha variant, while the earlier surges were dictated by the original SARS-CoV-2 virus ^7^. Analysis of RT-PCR data from Israel revealed an increase of 40% in transmission for the alpha SARS-CoV-2 variant relative to the Wildtype strain ^7^. This period coincided with the Israeli vaccination campaign, which included offering the Pfizer mRNA BNT162b2 vaccine to pregnant women due to the increasing evidence of severe and compromising COVID-19 complications during pregnancy ^8,9^, together with the indication that the Alpha variant poised increased severity and mortality for women relative to men ^10^.

A recent study highlighted the fact that pregnant women have a reduced antibody response against SARS-CoV-2 when compared to non-pregnant women, with lower titers of IgG-S-RBD ^11^. Studies by us and others demonstrated the safety and benefits of vaccination during pregnancy, promoting its implementation for preventive maternal-fetal care ^12,13^. The humoral response to SARS-CoV-2 infection and vaccination in pregnancy showed that IgG efficiently crosses the placenta providing immunity for the newborn ^13-19^. However, little is known about maternal and neonate IgG concentrations at term following early pregnancy infection or vaccination, as well as the impact of infection by VOC.

In contrast to IgG, IgM antibodies do not cross the placental barrier thereby providing a sensitive measure for fetal exposure to viral antigens or the virus itself. Cases of cord IgM positivity following SARS-CoV-2 infection during pregnancy are relatively rare but have been reported by us and others ^13,19^, suggesting potential placental damage, exposure of the fetus to viral antigens, or in rare cases, vertical transmission ^20^. As opposed to infection, no cord blood IgM for spike antigens has been detected among vaccinations ^13^, consistent with no fetal exposure to antigens derived from the vaccine.

Herein, we report updated findings from the Israel Covid-19 in Pregnancy study ^13^, concerning the evolution and dynamics of the maternal-fetal humoral response to SARS-CoV-2 infection and vaccination with the pandemic progression. Specifically, this work addresses for the first time the humoral response and waning of immunity for infection and vaccination during pregnancy, comparing the impact of the Wildtype versus Alpha variant dominated periods in Israel (2^nd^ versus 3^rd^ waves respectively).

## Materials and Methods

### Study Design

Pregnant women admitted for delivery at eight medical centers in Israel (Hadassah Mount-Scopus, Wolfson, HaEmek, Hillel Yafe, Rabin, Shaare Zedek, Meir, and Sourasky Medical Centers) were approached for enrollment in the Israel Covid-19 in Pregnancy study, starting in April 2020. Eligibility criteria included maternal age ≥18 years and willing to provide informed consent. Pregnant women with active maternal COVID-19 disease were excluded from this study. The project cohort consisted of 1702 participants which were initially stratified into three main groups: 427 vaccine recipients (all collected between January to June 2021), 211 participants with past SARS-CoV-2 infection during pregnancy, and 1039 unvaccinated women without prior documentation for infection (Figure 1a).

**Figure 1:**
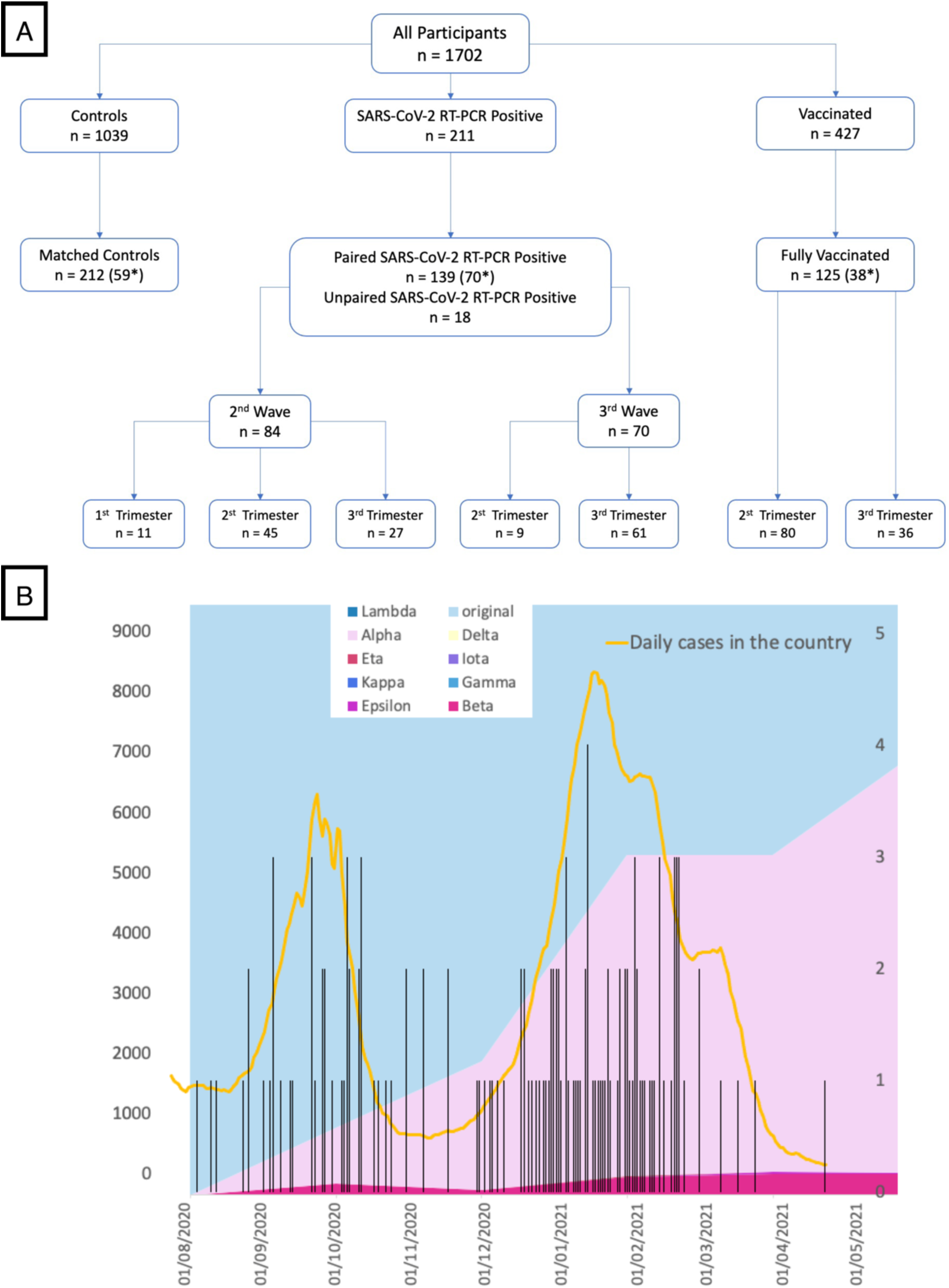
Study Design. **(A)** Patients were recruited from 8 Medical Centers in Israel and were all SARS-CoV-2 negative at delivery. Unpaired samples (maternal serum only, n=18) were included in the SARS-CoV-2 group. The SARS-CoV-2 wave groups were further classified based on the gestational age (GA) of infection: GA < 13 for 1^st^ trimester, 13 ≤ GA < 27 for 2^nd^ trimester, and 27 ≤ GA for 3^rd^ trimester. Vaccinated participants were also classified into two groups according to the GA of their first vaccine shot, 2^nd^ trimester (13 ≤ GA < 27) and 3^rd^ trimester (27 ≤ GA). * Asterisk marks the number of previously published samples ^13^. **(B)** Variants (background colors: light blue = original Wildtype virus, pink = Alpha variant), daily SARS-CoV-2 positive cases in Israel (yellow line, left y-axis) showing the 2^nd^ and 3^rd^ waves, and SARS-CoV-2 positive cases in our study (black bars, right y-axis). The variants’ data was obtained from ‘CoVariants’ ^33^ and the Israeli infection incidence from ‘Our Worls in Data’ ^34^.

Out of the 211 past SARS-CoV-2 positive participants, 139 maternal-fetal matched and 18 maternal only sera samples were included in the analyses (including 70 reported in our previous study ^13^). The 157 PCR-positive samples were divided into two groups based on their PCR-positive date (“date of infection”), representing the SARS-CoV-2 infection waves observed in Israel (Figure 1b). Samples were allocated to the 2^nd^ wave if their PCR-positive date was before the 1^st^ of January 2021 (n=84) and to the 3^rd^ wave if they got infected afterwards (n=70), based on the timing of 50% prevalence of the alpha variant in Israel ^7^. Among the 427 vaccinated participants, 125 fully vaccinated paired samples were included (38 from our previous study ^13^). Lastly, a control sub-group (n=212) was selected: 59 were reported in a previous publication from this project ^13^, and the remaining 153 were matched to the SARS-CoV-2 positive group based on clinical parameters (Table 1). Overall, serology results were obtained for 494 Dyads: n=157 with past RT-PCR positive results, n=125 fully vaccinated and n=212 controls (Figure 1a). Participant clinical and neonate outcome characteristics are provided by study groups in Table 1.

**Table 1.** Participants clinical and neonate characteristics.

|  | **Control group**<br>**n = 212** | **Past SARS-Cov-2 group**<br>**n = 157** | **Vaccinated group**<br>**n = 125** |
| --- | --- | --- | --- |
| Maternal age, mean ± SD , years | 29.5 ± .55<sup>a</sup> | 28.7 ± 5.5<sup>a</sup> | **31.4 ± 6.1<sup>b</sup>** |
| Gestational age, mean ± SD, weeks | 39 ± 1.5* | 39 ± 1.7 | **39 ± 1.6\*** |
| Preterm delivery (<37), n (%) | 18 (8.5) | 14 (8.9) | **10 (8)** |
| Pregravid BMI (Kg/m<sup>2</sup>), mean ± SD | 24.6 ± 5.5*<br>n = 184 | 25.3 ± 5.6<br>n = 142 | **24.4 ± 4.7\***<br>**n = 114** |
| Gravidity, median (IQR) | 2 (3)<sup>a</sup> | 3 (2)<sup>a,b</sup> | **3 (3)<sup>b</sup>** |
| Parity, median (IQR) | 1 (2)<sup>a</sup> | 2 (2)<sup>b</sup> | **1 (2)<sup>a,b</sup>** |
| Maternal comorbidities, n (%) |  |  |  |
| Hypertensive disorders | 8 (3.8) | 4 (2.5) | **5 (4)** |
| Diabetes or gestational diabetes | 14 (6.6) | 5 (3.2) | **9 (7.2)** |
| Asthma | 3 (0.5) | 4 (2.5) | **1 (0.8)** |
| Thyroid disease | 9 (4.2) | 5 (3.2) | **8 (6.4)** |
| Smoker | 9 (4.2) | 2 (1.3) | **5 (4)** |
| Infant sex, n (%) |  |  |  |
| Male | 108 (50.9)<sup>a</sup> | 79 (50.3)<sup>a,b</sup> | **48 (38.4)<sup>b</sup>** |
| Female | 104 (49.1)<sup>a</sup> | 78 (49.7)<sup>a,b</sup> | **77 (61.6)<sup>b</sup>** |
| Birthweight, mean ± SD, grams | 3215.8 ± 464.2 | 3301.3 ± 506.4 | **3252.7 ± 472.2** |
| NICU, n(%) | 11 (6.5) | 6 (4.9) | **4 (5.8)** |
| PCR- positivity GA, mean ± SD, weeks | - | 26.3 ± 8.3 | - |
| First vaccine dose GA, mean ± SD,<br>weeks | - | - | **27 ± 5.4** |

Continuous parameters were analyzed by Kruskal-Wallis one-way ANOVA test, following by Dunn's all-pairwise comparisons tests; Pearson Chi-square analysis was used to compare proportional data. <sup>a,b</sup> within a row, values without a common superscript significantly differed ( $P < 0.05$ ). \* within a row marks differences in the recruited groups.

### Samples Collection and Handling

Maternal and cord serum samples were collected from the enrolled patients prior to delivery, and from the umbilical cord just following delivery, and handled as detailed in the Supplementary Material ^13^.

### Quantification of Antibodies

Serum samples were prepared and analyzed using multiplexed fluorescent bead assay as previously described (details in Supplementary Material ^13^). Briefly, samples were thawed, heat-inactivated at 56 °C for 30 minutes, and transferred to bar-coded 96-well plates for analysis. Serum IgG and IgM were detected using Milliplex MAP SARS-CoV-2 Antigen Panel 1 IgG (HC12SERG-85K) and IgM (HC19SERM1-85K) supplemented with additional HA-coupled microspheres. Recombinant 6XHIS-HA protein derived from two isolates of influenza (Phuket /Singapore) were purchased from Immune Tech Corp and coupled to Bioplex Mag COOH beads according to manufacturer’s instructions (Bio-Rad). Coupling was confirmed by incubating with 1:1000 anti-HIS-PE (abcam) for 1 hour and washing 3 times with Milliplex wash buffer, with detection in a MAGPIX reader. Positive and negative controls were included in each analysis for accuracy and reproducibility.

### Statistical Analyses

Statistical analyses were performed using Statistix 8 software (Analytical Software, Tallahassee, FL USA) and Prism 5.01 (GraphPad Software; San-Diego, CA, USA). IgG and IgM antibody (S1, S2, RBD, N) MFI were log10-transformed for the analyses. Comparisons of antibody concentrations and other continuous parameters (*e*.*g*., clinical data, transfer ratios) among groups were analyzed by Kruskal– Wallis one-way ANOVA test, following by Dunn’s all-pairwise comparisons test; or alternatively, by Wilcoxon Rank Sum Test (if only two groups were compared). Additionally, when relevant, the effects of wave (either the 2^nd^ or 3^rd^ wave), trimester (1^st^, 2^nd^, 3^rd^), and the wave by trimester interaction on IgG and IgM antibody concentrations or their transfer ratios were analyzed in a General Analysis of Variance Test (ANOVA), followed by Tukey HSD All-Pairwise Comparisons test. The equality of variances of maternal and cord blood antibody concentrations among groups (Control; Vaccinated; PCR-positive 2^nd^ wave; PCR-positive 3^rd^ wave) were evaluated by Levene’s Test, O’Brien’s Test, as well as by Brown and Forsythe Test. Pearson Chi-square analysis was used to compare proportional data. Pearson’s Correlation and Linear Regression tests were used to determine various correlations between maternal and cord blood IgG and IgM antibodies, as well as to test associations between time of exposure (*i*.*e*., to SARS-CoV-2 virus or to the Vaccine) and IgG and IgM concentrations. All statistical tests were based on two-tailed hypotheses. Differences were considered significant at P < 0.05.

## Results

### Participant Characteristics

Analysis of the study groups showed differences in maternal age, gravidity, parity and infant sex between the vaccinated and the control groups (Table 1). Within the control group, 35 participants (16.5%) were found to be seropositive for N using IgG-N (mean net fluorescence intensity (MFI) > 1583) as the threshold ^13^, which together with IgG seropositivity to the other SARS-CoV-2 antigens (S1, S2 and RBD), is indicative of induced immunity due to infection. Similarly, 17 vaccinated participants (13.6%) were seropositive for IgG-N leading to an overall undiagnosed seropositivity rate of 15.43%. High fetal IgM response was defined as IgM-N > mean + 2SD (MFI > 741), and cord serum seropositivity was set for samples having a high IgM response for at least two antigens. Accordingly, 17 neonates (3.6 %): 7 from the control, 6 from the infected and 4 from the vaccinated groups were found to have a high IgM response (Figure S1).

### Maternal and Fetal Humoral Response Across Gestation

Maternal and cord serological response to SARS-CoV-2 infection were analyzed by trimester and wave of infection (Figure 2 and 3, Table S1 and S2). The general AOV/AOCV test (Table S2) revealed an interaction between the wave and trimester of infection for all maternal IgG antigens, IgM-S2 and IgM-N (P<0.05), excluding variant induced changes in affinity to the assay antigen as the prime impact. A gradual decline in maternal IgG and IgM concentrations could be detected with increased interval between infection and birth. Specifically, in the second wave, MFI of maternal IgG-S1, IgG-RBD, IgG-N, IgM-S1 and cord blood IgM-S1 were significantly lower at birth in women infected on the 1^st^ or 2^nd^ trimester, as compared to those infected in the 3^rd^ trimester (P<0.05). Similarly, maternal IgG for all antigens, IgM-S2 and IgM-N were significantly lower in women infected on the 2^nd^ trimester during the third wave, as compared to those infected in the 3^rd^ trimester (P<0.05). Furthermore, maternal MFI for all IgG antigens, IgM-S2 and IgM-N were significantly higher for 2^nd^ trimester infections during the 2^nd^wave as compared to the 3^rd^ wave (P<0.05). Additionally, maternal IgM-S1 and IgM-RBD were significantly higher for 3^rd^ trimester infections during the 2^nd^ wave, as compared to 3^rd^ trimester infections during the 3^rd^ wave (P<0.05). Interestingly, cord IgM-S1 and IgM-S2 were significantly higher for 3^rd^ trimester infections during the 3^rd^ wave, as compared to infections during the 2^nd^ wave (P<0.05).

**Figure 2:**
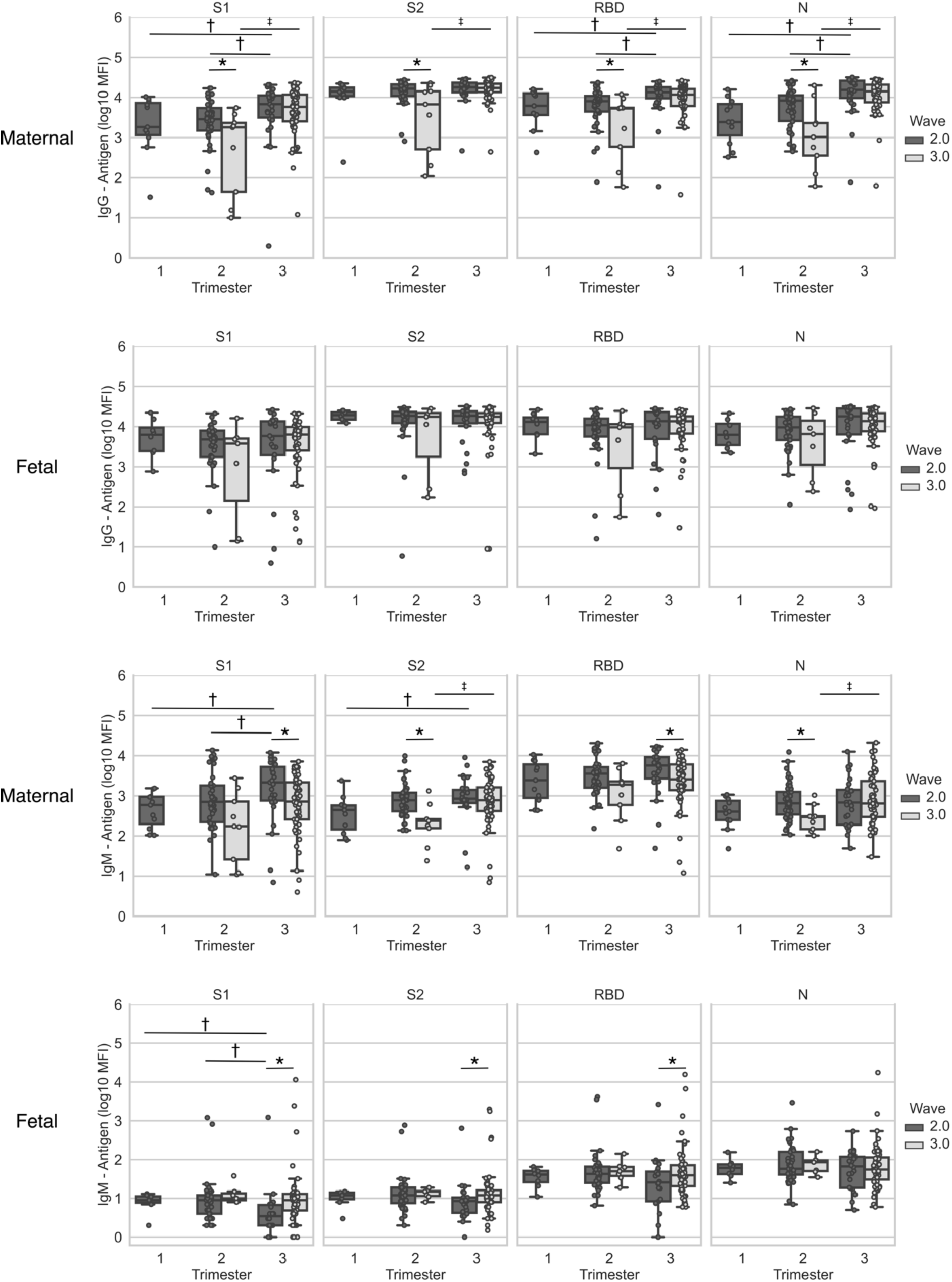
Maternal and fetal humoral response to SARS-CoV-2 across gestation. Maternal and cord blood was derived at the time of delivery from patients who recovered from infection during pregnancy with SARS-CoV-2 verified by RT-PCR (n=161). Maternal and fetal IgG and IgM antigens (S1, S2, RBD and N) concentrations are plotted per wave and per trimester of infection. Top two rows, IgG; bottom two rows, IgM. Statistical significance: * indicate significant differences between waves of infection within the same trimester (Wilcoxon Rank Sum Test), † indicate differences between trimesters in the 2^nd^ wave (Kruskall–Wallis one-way ANOVA), and ‡ indicates differences between trimesters in the 3^rd^ wave (Wilcoxon Rank Sum Test). Box and whiskers: Middle line = Median; Box= 25^th^ and 75^th^ percentiles; whiskers= are the minimum (Q1 – 1.5*IQR) and maximum (Q3 + 1.5*IQR).

**Figure 3:**
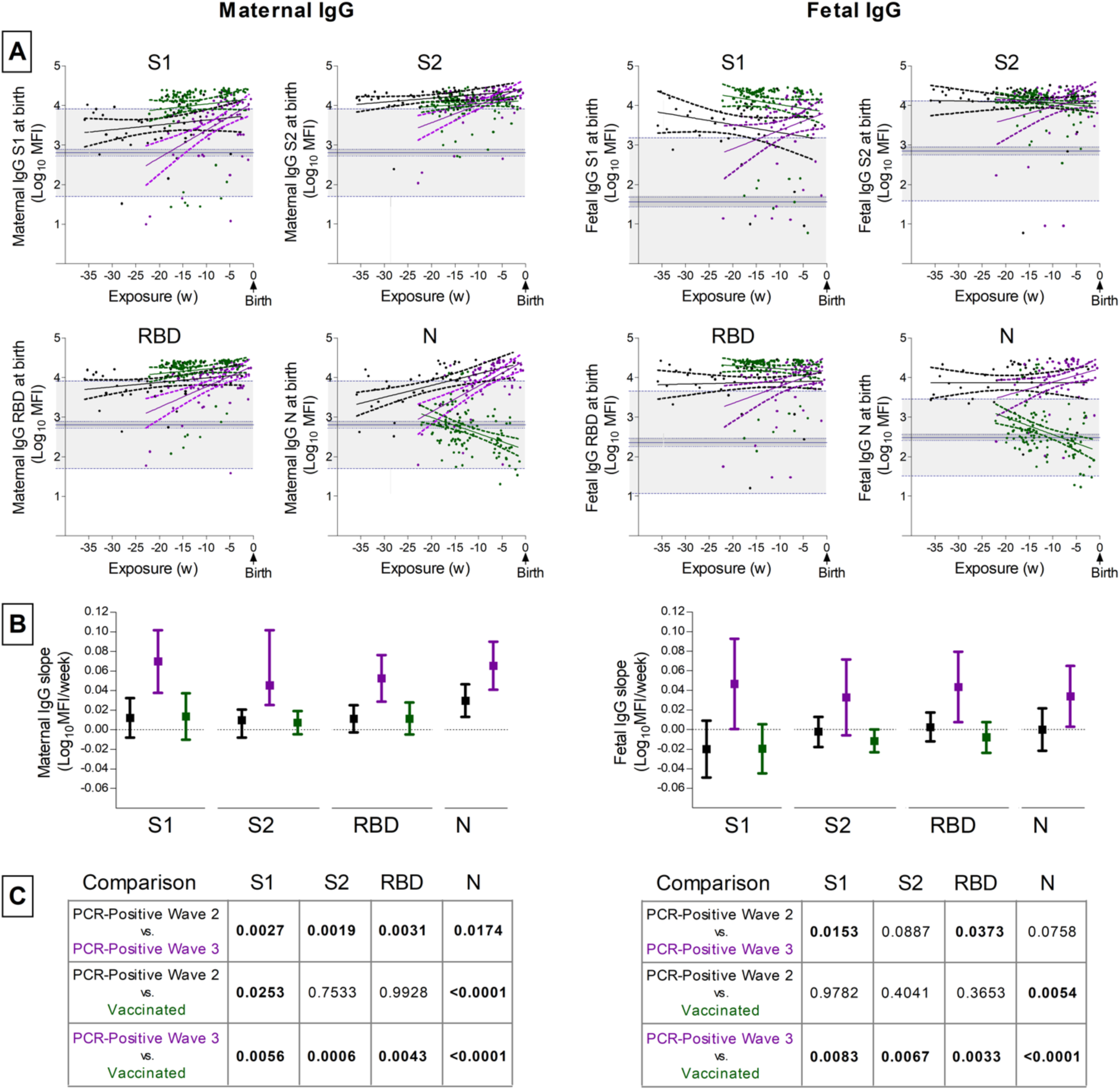
SARS-CoV-2 Antibody Level Decay Across Gestation. **(A)** IgG maternal (left column) and fetal (right column) antibody concentrations at the time of delivery are plotted (Y-axis) by the time of exposure, either to the viral antigens (i.e., positive-PCR) or to vaccine antigens, in relation to the time of delivery (x-axis). The X-axis represents the actual time from exposure to sampling. The data calculated from the true control group (defined as all seronegative samples, using IgG-N log10(MFI) < 3.19948 ^13^) is provided as Mean (straight line), 95% CI (dotted lines above and below, highlighted in dark gray) and Mean ± 2SD (highest and lowest dashed lines, highlighted in light gray) for reference purposes. Infections during the 2^nd^ wave are plotted in black, infections during the 3^rd^ wave in purple and vaccinations in green. **(B)** The mean antibody reduction rate (or calculated slope) was calculated using linear regression analyses, and is plotted by antigen and per group as a dot, together with its 95% CI (bars). **(C)** The slope of the antibody decay was compared among the groups. P values for each comparison are listed in the occupying tables and highlighted in bold where significant.

In the vaccinated group, the data were analyzed by the trimester of the first vaccine dose (Figure S2, Table S3). MFI of maternal IgG-S1, IgG-RBD and IgM-S1 were significantly higher at birth for 3^rd^ trimester vaccination (P<0.05) as compared to 2^nd^ trimester vaccination.

The stability of maternal and fetal humoral protection induced by infection or vaccination was evaluated by analysis of the interval between exposure and delivery, thereby accounting for variation in gestational age at delivery (Figure 3a). Similar decay of antibody concentration was observed between vaccination and 2^nd^ wave infections (Figure 3b). However, decay of maternal and fetal IgG for 3^rd^ wave infection was significantly faster than for 2^nd^ wave infection or vaccination (Figure 3b & 3c).

### Maternal to Fetal Antibody Transfer

IgG transfer ratio (TR) was calculated as cord divided by maternal MFI levels. TR of IgG-S1, IgG-S2 and IgG-RBD at delivery was significantly higher for 2^nd^ compared to 3^rd^ trimester vaccination. Similarly, IgG-N TR was higher for 2^nd^ versus 3^rd^ trimester infections during the 2^nd^ wave (P<0.05). Differences were also found between vaccinated and 2^nd^ wave infections for both 2^nd^ and 3^rd^ trimester exposure (Figure S3, Table S4, P<0.05).

The impact of SARS-CoV-2 infections on the maternal-fetal IgG transfer was analyzed for equality of variances using three statistical tests (Table S5). The paired maternal-fetal serological responses are plotted by groups in Figure 4, together with data density curves. The data shows no significant differences for maternal IgG-HA(Phuket) and IgG-HA(Singapore) MFI variances between the groups. Surprisingly, cord IgG-HA(Phuket) and IgG-HA(Singapore) MFI variances were significantly higher for 2^nd^ and 3^rd^ wave infections as compared to the vaccinated and control groups (Table S5). On the other hand, maternal and cord SARS-CoV-2 antibody level variances differed only between the control and the remaining groups (Table S5). No sexual dimorphism in IgG was observed for any of the measured parameters. Namely maternal and cord IgG levels were similar for male and female neonates (Figure S4)

**Figure 4:**
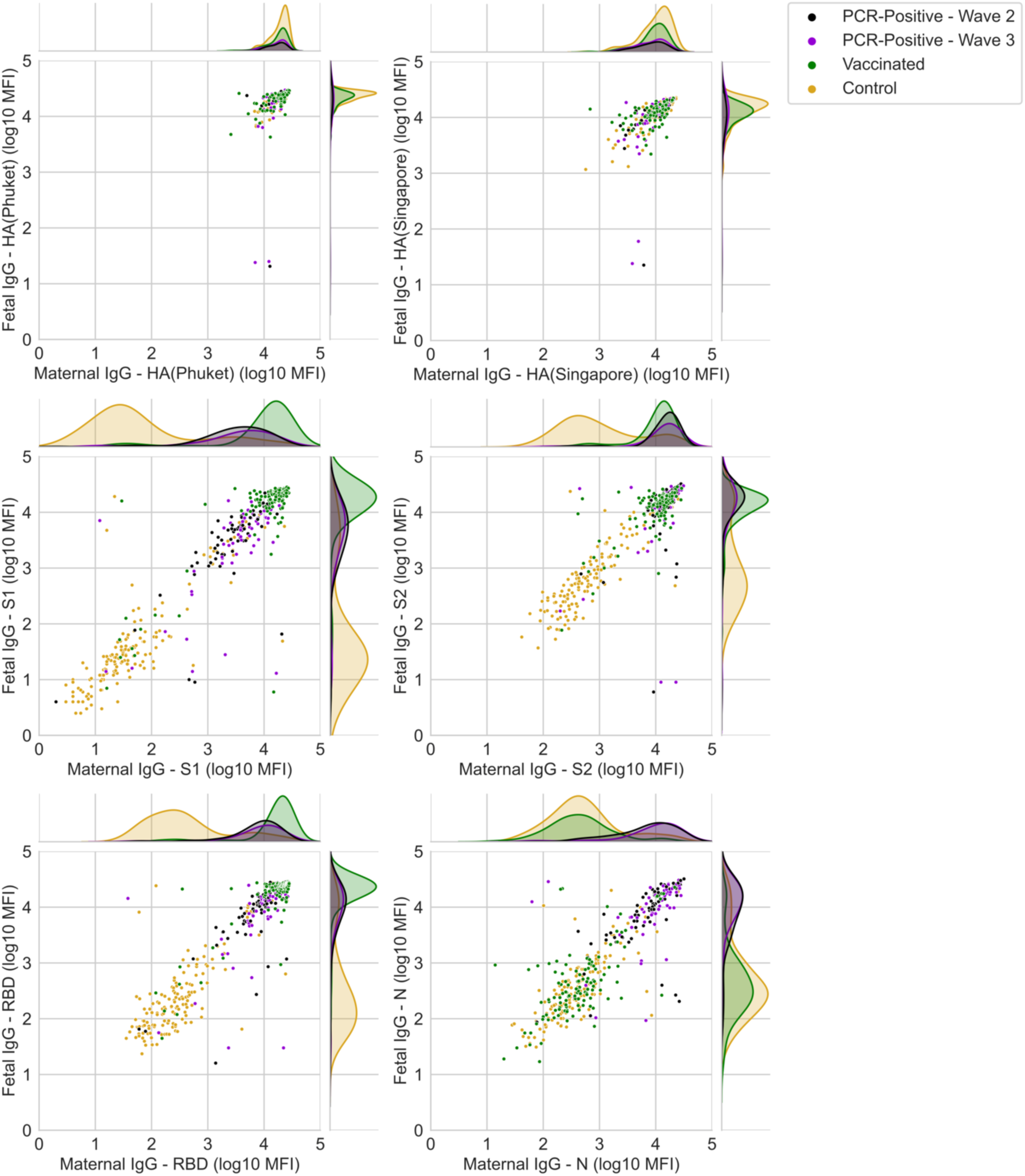
Waves Effect on IgG Transfer. Maternal and cord serological response for HA(Phuket), HA(Singapore), S1, S2, RBD and N plotted for the SARS-CoV-2 group, divided according to the waves of infection, vaccinated and control groups. Each dot represents data from a single patient and the marginal are density curves. The equality of variances of maternal and cord blood antibody concentrations among groups were evaluated by Levene, O’Brien, and Brown and Forsythe tests as detailed in Supplementary Table S5.

### Cord Serum IgM

Among the 476 cord samples tested, 17 samples were identified with a robust IgM response to at least three SARS-CoV-2 IgM antigens. Figure S5 highlights the 17 samples having high IgM response (big dots), which cluster in the top right corner when plotting IgM-RBD against IgM-N (second row, right column); it can be observed that contrary to the IgG response (top row), the IgM responses are independent from the maternal serological response (second row, left column). Additionally, the cord serum positive samples showed also a high response to IgM-HA(Phuket) and IgM-HA(Singapore) (bottom row, right column).

## Discussion

Pregnant women have been at an increased risk of complications since the beginning of the coronavirus pandemic. Moreover, pregnant women have more severe infections and worse pregnancy outcomes for those infected during the SARS-CoV-2 Alpha variant dominated period compared to Wildtype infections and to non-pregnant women ^21^, but also have reduced IgG-S-RBD titers as compared to non-pregnant women ^11^. We report here for the first time that the maternal humoral response to SARS-CoV-2 during the Alpha variant dominated period is significantly weaker when compared to infections during the Wildtype virus dominated wave. These findings suggest that the SARS-CoV-2 Alpha variant is less immunogenic, and perhaps can partially evade the immune system of pregnant women.

Cord serum anti-SARS-CoV-2 IgG concentrations were similar for infections in the 2^nd^ and 3^rd^ waves, indicating that the transfer of immunity to the developing fetus is less sensitive to waning of maternal IgG and less susceptible to SARS-CoV-2 mutations. These results highlight the importance of placental regulation and active transport for fetal immunity. Interestingly, the variance of cord serum IgG-HA(Phuket) and IgG-HA(Singapore) concentrations was significantly higher for the PCR-positive group relative to control and vaccinated despite the fact that variances among maternal levels were similar. These results implicate SARS-CoV-2 infection during pregnancy in disruption of placenta IgG, resulting in a decreased neonate immunity against Influenza. This is is consistent with recent reports of placental SARS-CoV-2 infections and damage ^22-24^. Lastly, in contrast to a recent study of a smaller cohort ^25^, we show here that cord IgG levels were similar for male and female neonates.

The decline of antibody titers following infection attracted much of attention because of the emergence of several variants of concern and since it serves as the basis for vaccination policies in previously infected or vaccinated individuals ^26^. A recent study showed that the decay rates for IgG-RBD-S in the general population were not significantly different between SARS-CoV-2 infection and vaccination, and that antibody titers were still detectable 6 months after exposure to antigens ^26^. Our data shows that in pregnant women the antibody decay rate is also similar for vaccinated and for infections during the 2^nd^ wave, dominated by the Wildtype variant. However, the decay rate was significantly faster for 3^rd^ wave infections. Together with the lower antibody titers found for infections during the Alpha variant dominated period, this study demonstrates that pregnant women mount a weaker and shorter immune response against SARS-CoV-2 Alpha variant compared to Wildtype infections.

Clinically, the results reported here underscore for the importance of vaccinating pregnant women, who were infected prior to pregnancy or during early stages of pregnancy, particularly with the SARS-CoV-2 alpha variant. Due to the rapid decay of antibodies, these women might be at increased risk for re-infection. Our understanding on the decline of mRNA BNT162b2 vaccine induced antibody titers across gestation remains poor. Evidence is needed to support policies of immunity boosting during the pre-natal period. In the present study, we demonstrate that following vaccination, there is a mild decline in maternal antibody titers across gestation, but not in cord serum antibody levels, supporting the efficiency of vaccination during pregnancy for neonate immunity. Therefore, vaccination during the second trimester provides defense to both mother and fetus. Interestingly, a high IgG transfer ratio was found for first trimester wave 2 infections indicating that infection in the early stages of pregnancy also leads to sustained fetal immunity.

Transplacental IgG transfer is mediated by FcRn, found in the fetal endothelium and within the syncytiotrophoblast layer, which is in direct contact with maternal blood. Ex vivo models have demonstrated that IgG bound to viral antigens can be transported to fetal blood ^27^, potentially triggering a fetal immune response. It is well known that there is no transport of IgM across the placenta; however, fetal IgM has been detected in fetal blood from about 13 weeks of gestation ^28^. Therefore, high fetal IgM concentrations may suggest an intrauterine infection or immunological stimulation ^29-31^ by viral antigens transported bound to IgG ^27^ or cross reactivity of low affinity fetal IgM. Cases of possible vertical SARS-CoV-2 transmission are rare, but have been previously reported ^13,20,32^. Hence, the robust IgM response to all SARS-CoV-2 antigens found in seventeen neonates could be a result of immunological stimulation and not necessarily a vertical transmission of the SARS-CoV-2 virus. The elevation of IgM for HA in these neonates is consistent with transplacental antigen presentation or placental dysfunction.

The present work possesses several limitations, including bias in sample collection due to daytime recruitment of participants, thereby excluding most emergency cases. The period of sample collection differed among groups, as the Israeli vaccination campaign started after the commencement of the study. Additionally, antibody levels were quantified using antigens of the original virus, and may thus show reduced affinity to antibodies induced by the alpha variant.

Pregnancy induces particular changes in a women’s immune system placing them at a vulnerable position when it comes to infections. Therefore, general population immunity dynamics are not always applicable to pregnant women. This is the first study to compare the evolution and dynamics of the maternal-fetal humoral response between Alpha variant and Wildtype SARS-CoV-2 infections, demonstrating that pregnant women response is variant dependent. Additionally, this work enhances the important role of active transport and placental regulation for fetal immunity supporting vaccination during pregnancy.

## Data Availability

All data produced in the present study will be available after peer reviewed publication, upon reasonable request to the authors

## Acknowledgement

This work was supported by an Israeli Science Foundation KillCorona grant 3777/19 (to MN, MK, SY, AM) and by a research grant from the Weizmann Institute Fondazione Henry Krenter (to MN). We would like to thank the patients who made this research possible.

## Conflict of Interest

The authors have declared that no conflict of interest exists.

## Tables and Figure Legends

**Table 1: Clinical parameters of the study groups**.

Clinical parameters of the study groups. The control sub-group was matched to the SARS-CoV-2 positive group based on maternal age, gestational age, BMI, gravidity, parity and maternal comorbidities. ^a,b^ Values without a common superscript and within a row differed between the study groups (P < 0.01).

## Notes

### Competing Interest Statement

The authors have declared no competing interest.

### Funding Statement

Israel Science Foundation (ISF) and by The Weizmann Institute Fondazione Henry Krenter

### Author Declarations

Helsinki Ethics Committee of the Wolfson Medical Center, as well as all committees of all Medical Centers and the IRB of the Weizmann Institute gave ethical approval for this work

