## Supplemental material for "Waning of the Humoral Response to SARS-CoV-2 in Pregnancy is Variant-Dependent"

### Supplementary Material

### Materials and Methods

#### Samples Collection and Handling

Serum dyads (maternal and fetal) were collected at delivery. Patients were enrolled and maternal blood was collected prior to delivery and from the umbilical cord following delivery. The umbilical cord was wiped clean and blood was drawn from the vein. Blood samples were centrifuged (1000*g*, 10 minutes at room temperature), and serum aliquotes were stored stored at –80°C in dedicated precoded tubes for analyses at the Weizmann Institute.

#### Quantification of Antibodies

Serum IgG and IgM were detected using Milliplex MAP SARS-CoV-2 Antigen Panel 1 IgG (HC12SERG-85K) and IgM (HC19SERM1-85K) supplemented with additional HA-coupled microspheres. Reagents were prepared according to manufacturer instructions and dispensed to 96-well source plates (Greiner 651201, Sigma-Aldrich). Serum samples were diluted 1:100 in assay buffer and added to antigen-immobilized Milliplex beads in 96-well plates using a Bravo liquid handler (Agilent). Plates were covered, shook for two hours at room temperature, and washed three times with wash buffer, using a manual magnet and multidrop combi dispenser (Thermo). Anti-IgG-PE or Anti-IgM-PE 16 conjugate was added, and the samples were incubated (90 minutes with shaking) and washed. Sheath fluid was added to the samples, and net fluorescent intensity (MFI) signals were detected on a Luminex MAGPIX reader Repeat measurements of the same sample showed less than 5% difference for all antibodies.

### Tables & Figures

**Supplementary Figure 1.** Serological heat maps generated from the acquired IgG and IgM data, segregated by the main recruitment groups (top 3 rows: control, PCR Positive, Vaccinated) and by the waves of infection for the SARS-CoV-2 group (bottom 3 rows: 2nd wave, 3rd wave). From left to right: Maternal IgG (S1, S2, RBD, N), Maternal IgM (S1, S2, RBD, N), Fetal IgG (S1, S2, RBD, N), Fetal IgM (S1, S2, RBD, N). Each row represents matched maternal-fetal data, ranked by the maternal IgG reactivity to the N antigen within each group (low, dark blue; high, yellow). Note the serologically positive mothers (high IgG-N in yellow) within the control and vaccinated groups, and the high IgM fetal responses.

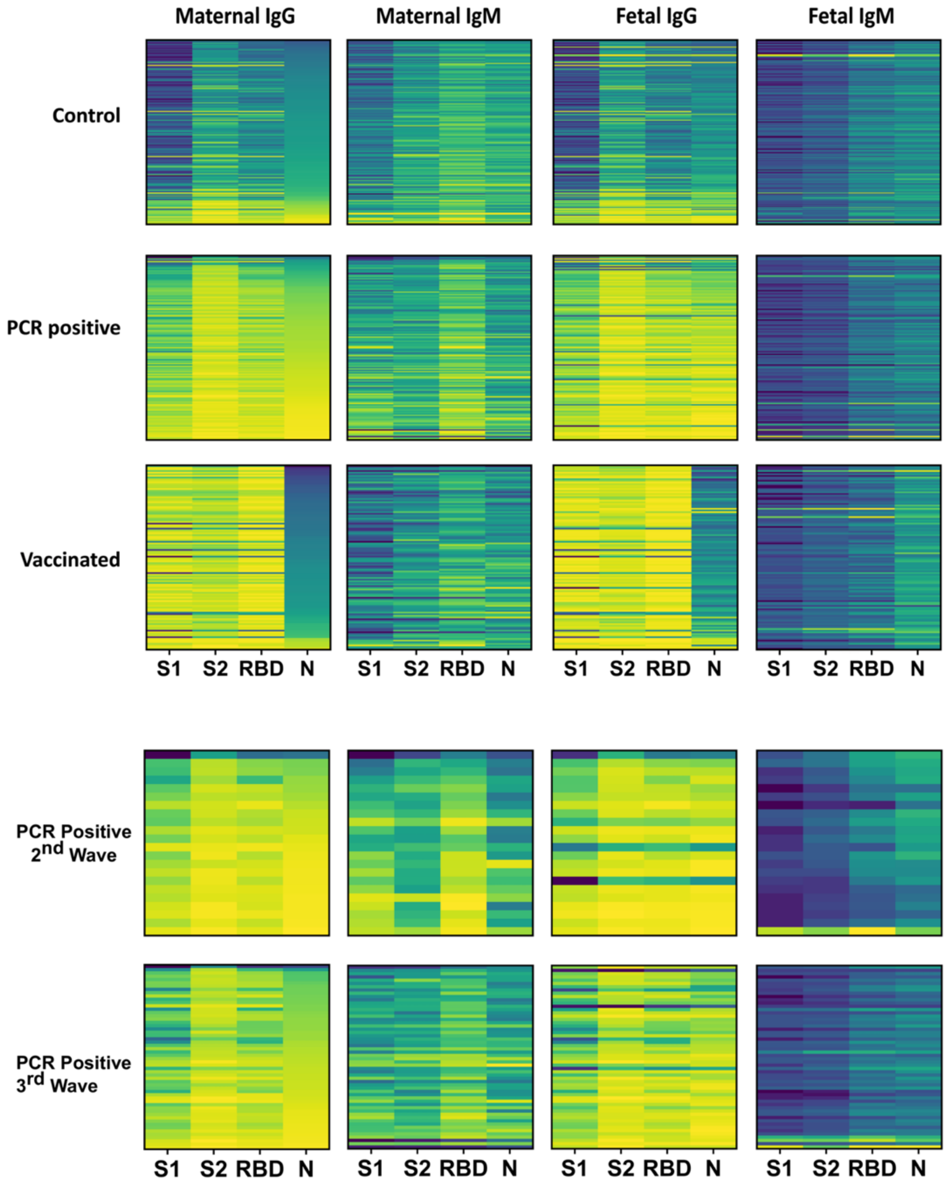

**Supplementary Figure 2.** Maternal and cord blood was derived at the time of delivery from patients who got fully vaccinated during pregnancy. Maternal and fetal IgG and IgM antigens (S1, S2, RBD and N) concentrations are plotted by the trimester of the first dose. Top two rows, maternal titers; bottom two rows, cord titers; left columns, IgG; right columns, IgM. * Indicate significant differences between trimesters of vaccination (Wilcoxon Rank Sum Test). Box and whiskers: Middle line = Median; Box= 25th and 75th percentiles; whiskers= are the minimum (Q1 – 1.5*IQR) and maximum (Q3 + 1.5*IQR).

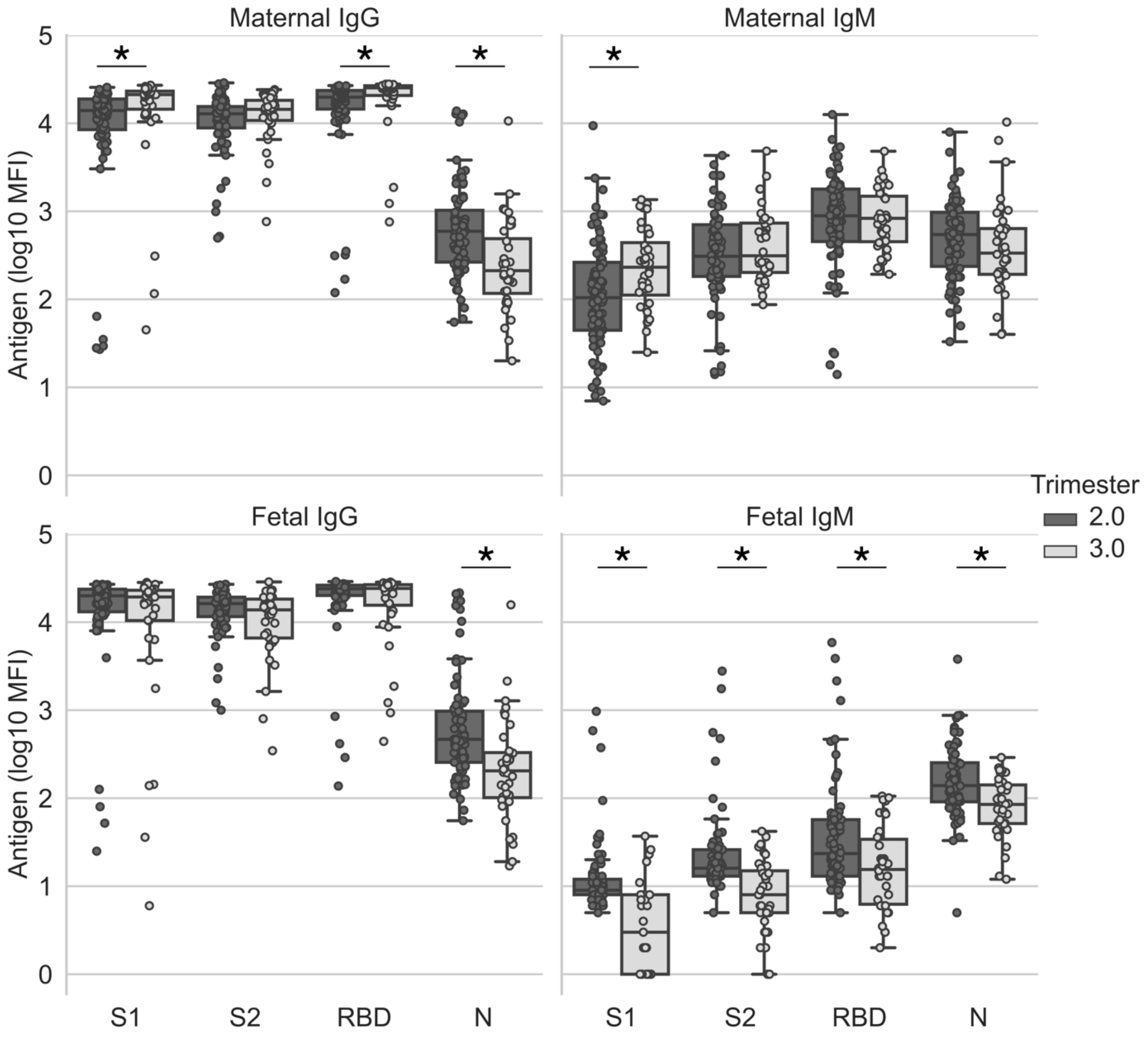

**Supplementary Figure 3.** Transfer ratio (cord IgG divided by maternal IgG levels) is plotted per wave and per trimester of infection or vaccination for all antigens (S1, S2, RBD and N). * Indicates significant differences between trimesters within the same group (Kruskall–Wallis one-way ANOVA), † indicates differences between infection/vaccination in the 2nd trimester (Wilcoxon Rank Sum Test), and ‡ indicates differences between infection/vaccination in the 3rd trimester (Wilcoxon Rank Sum Test). Box and whiskers: Middle line = Median; Box= 25th and 75th percentiles; whiskers= are the minimum (Q1 – 1.5*IQR) and maximum (Q3 + 1.5*IQR).‡

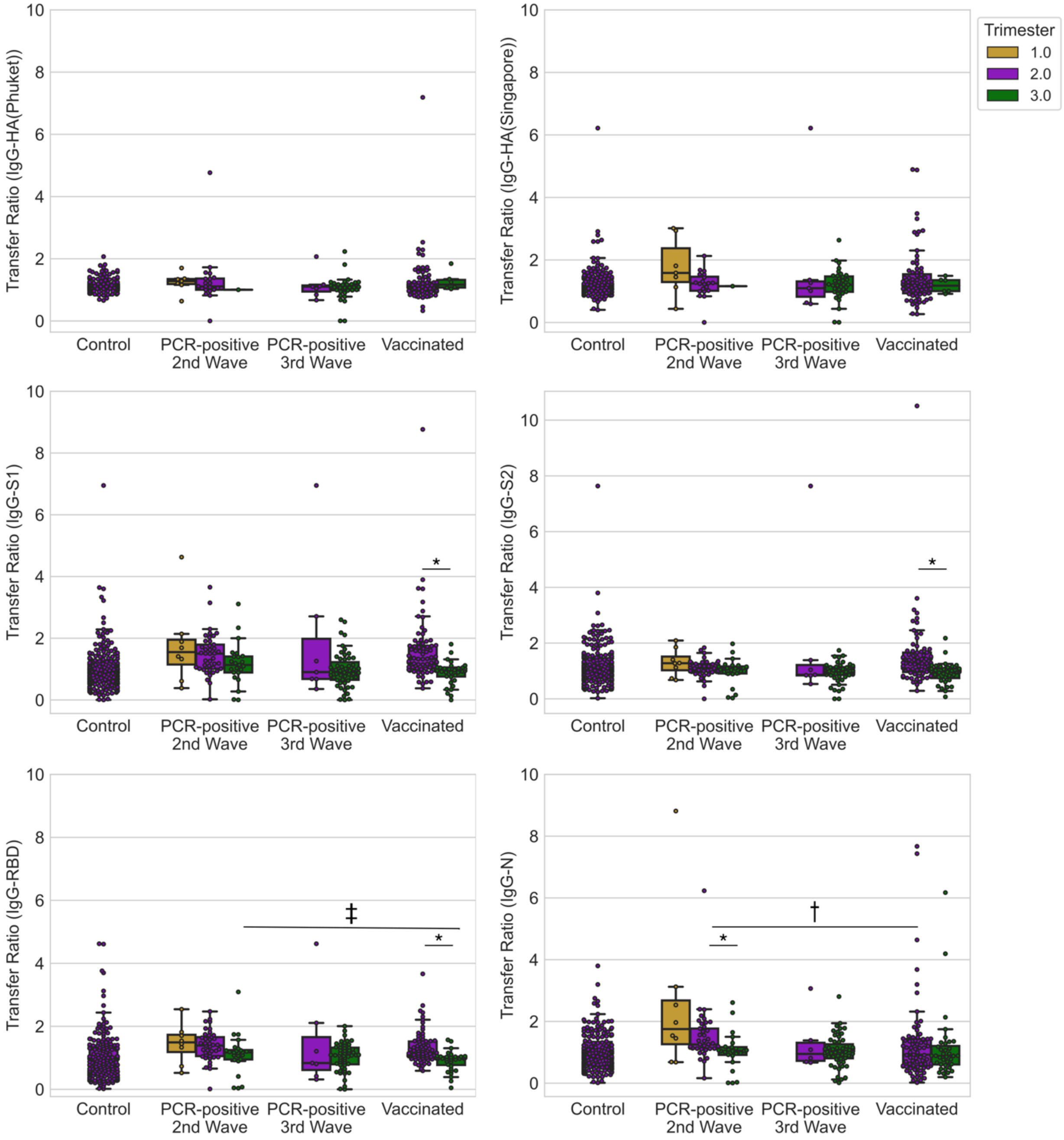

**Supplementary Figure 4.**

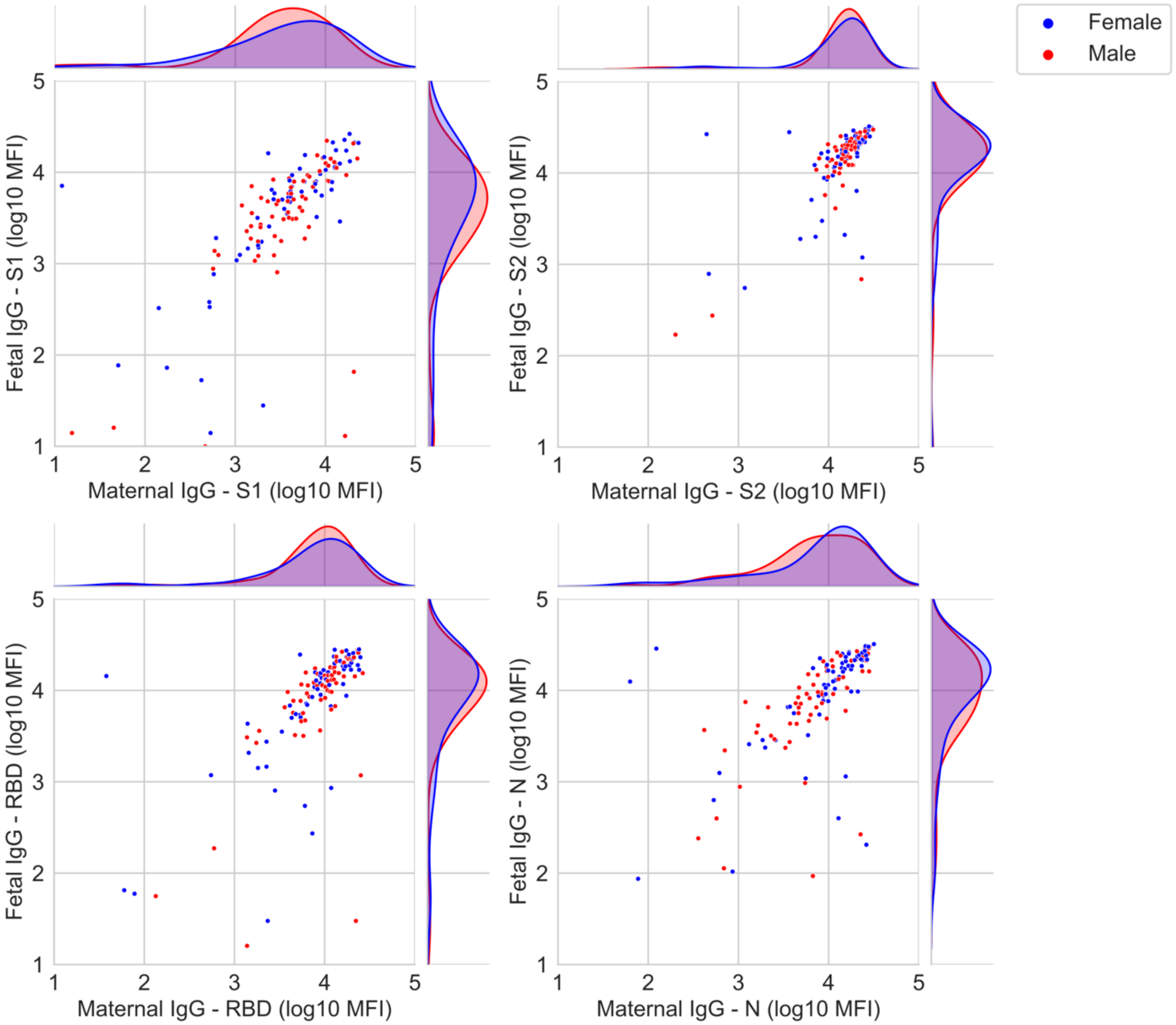

**Supplementary Figure 5.** Analysis of all study participants by maternal (left column) and cord (right column) serum IgG (top row) and IgM (2 bottom rows) response. By plotting IgG-N vs. IgG-RBD the data re-clusters by groups, where high IgG titer for N reveals patients exposed to the virus. Seventeen high cord IgM samples are highlighted by big dots, and cluster in the top right corner when plotting cord IgM-N vs. IgM-RBD. High IgM cord samples also showed high titers for IgM-HA(Phuket/Singapore) shown in the bottom row. Note that maternal and cord IgG plots mirror (top row), but IgM plots do not (bottom rows), showing the independent fetal response to antigens.

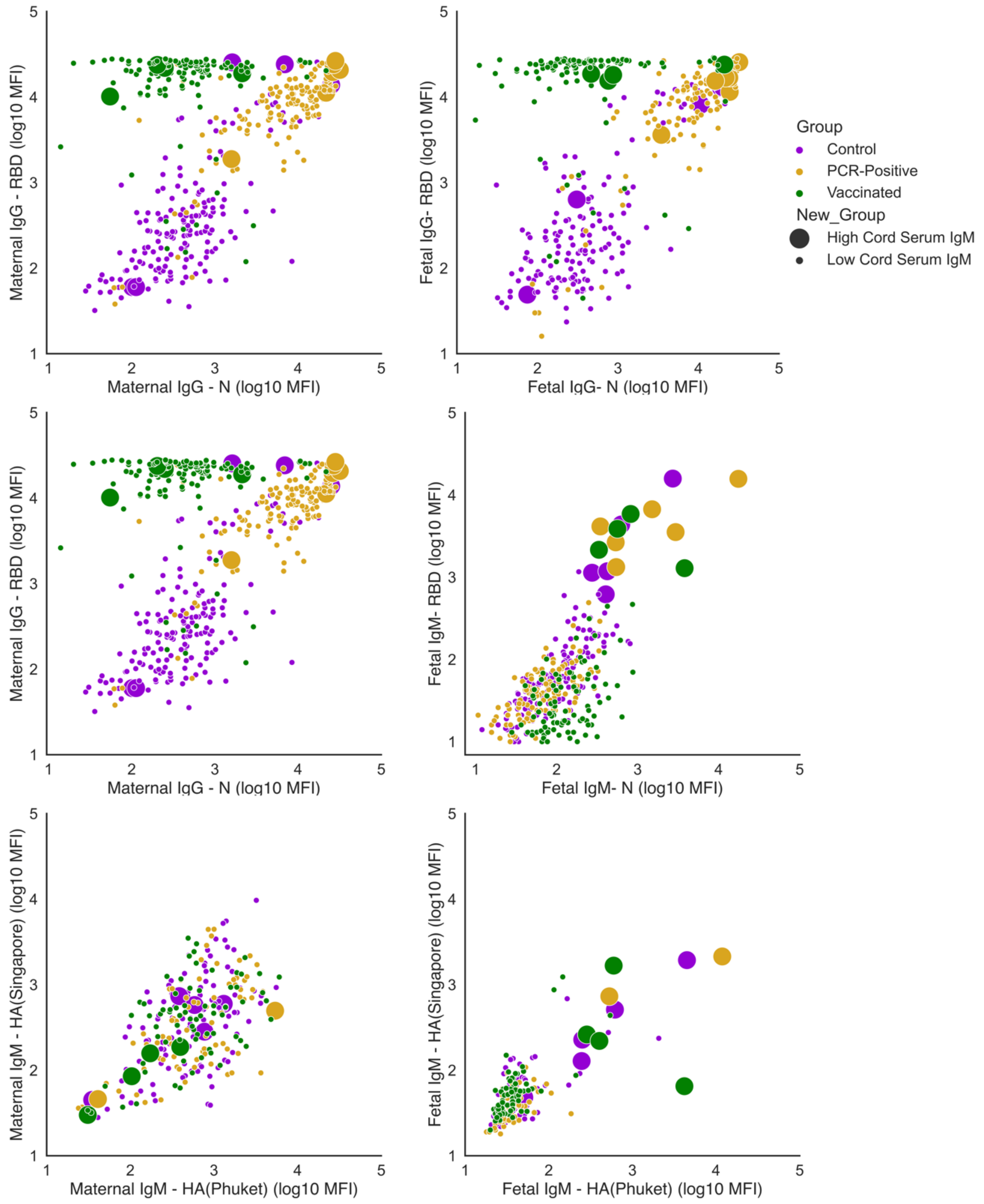

Tables

**Supplementary Table 1.** Statistical analysis of maternal and fetal serological response across gestation. Analysis of the data presented in Figure 2.

| Ab Type | Comparison | Statistical Test | P Value |
|---|---|---|---|
| **IgG – S1** | Maternal Ab concentrations between trimesters from infections during the 2nd wave | Kruskall–Wallis one-way ANOVA | P = 0.01 |
| | Fetal Ab concentrations between trimesters from infections during the 2nd wave | Kruskall–Wallis one-way ANOVA | P = 0.48 |
| | Maternal Ab concentrations between trimesters from infections during the 3rd wave | Wilcoxon Rank Sum Test | P = 0.001 |
| | Fetal Ab concentrations between trimesters from infections during the 3rd wave | Wilcoxon Rank Sum Test | P = 0.14 |
| | Maternal Ab concentrations between waves from 2nd trimester infections | Wilcoxon Rank Sum Test | P = 0.03 |
| | Fetal Ab concentrations between waves from 2nd trimester infections | Wilcoxon Rank Sum Test | P = 0.28 |
| | Maternal Ab concentrations between waves from 3rd trimester infections | Wilcoxon Rank Sum Test | P = 0.61 |
| | Fetal Ab concentrations between waves from 3rd trimester infections | Wilcoxon Rank Sum Test | P = 0.53 |
| **IgG – S2** | Maternal Ab concentrations between trimesters from infections during the 2nd wave | Kruskall–Wallis one-way ANOVA | P = 0.19 |
| | Fetal Ab concentrations between trimesters from infections during the 2nd wave | Kruskall–Wallis one-way ANOVA | P = 0.90 |
| | Maternal Ab concentrations between trimesters from infections during the 3rd wave | Wilcoxon Rank Sum Test | P = 0.009 |
| | Fetal Ab concentrations between trimesters from infections during the 3rd wave | Wilcoxon Rank Sum Test | P = 0.63 |
| | Maternal Ab concentrations between waves from 2nd trimester infections | Wilcoxon Rank Sum Test | P = 0.02 |
| | Fetal Ab concentrations between waves from 2nd trimester infections | Wilcoxon Rank Sum Test | P = 0.44 |
| | Maternal Ab concentrations between waves from 3rd trimester infections | Wilcoxon Rank Sum Test | P = 0.56 |
| | Fetal Ab concentrations between waves from 3rd trimester infections | Wilcoxon Rank Sum Test | P = 0.69 |

| | | | |
|---|---|---|---|
| **IgG –RBD** | Maternal Ab concentrations between trimesters from infections during the 2nd wave | Kruskall–Wallis one-way ANOVA | P = 0.001 |
| | Fetal Ab concentrations between trimesters from infections during the 2nd wave | Kruskall–Wallis one-way ANOVA | P = 0.31 |
| | Maternal Ab concentrations between trimesters from infections during the 3rd wave | Wilcoxon Rank Sum Test | P = 0.001 |
| | Fetal Ab concentrations between trimesters from infections during the 3rd wave | Wilcoxon Rank Sum Test | P = 0.15 |
| | Maternal Ab concentrations between waves from 2nd trimester infections | Wilcoxon Rank Sum Test | P = 0.04 |
| | Fetal Ab concentrations between waves from 2nd trimester infections | Wilcoxon Rank Sum Test | P = 0.35 |
| | Maternal Ab concentrations between waves from 3rd trimester infections | Wilcoxon Rank Sum Test | P = 0.22 |
| | Fetal Ab concentrations between waves from 3rd trimester infections | Wilcoxon Rank Sum Test | P = 0.39 |
| **IgG – N** | Maternal Ab concentrations between trimesters from infections during the 2nd wave | Kruskall–Wallis one-way ANOVA | P < 0.0001 |
| | Fetal Ab concentrations between trimesters from infections during the 2nd wave | Kruskall–Wallis one-way ANOVA | P = 0.06 |
| | Maternal Ab concentrations between trimesters from infections during the 3rd wave | Wilcoxon Rank Sum Test | P = 0.001 |
| | Fetal Ab concentrations between trimesters from infections during the 3rd wave | Wilcoxon Rank Sum Test | P = 0.16 |
| | Maternal Ab concentrations between waves from 2nd trimester infections | Wilcoxon Rank Sum Test | P = 0.01 |
| | Fetal Ab concentrations between waves from 2nd trimester infections | Wilcoxon Rank Sum Test | P = 0.46 |
| | Maternal Ab concentrations between waves from 3rd trimester infections | Wilcoxon Rank Sum Test | P = 0.13 |
| | Fetal Ab concentrations between waves from 3rd trimester infections | Wilcoxon Rank Sum Test | P = 0.25 |
| **Ab Type** | **Comparison** | **Statistical Test** | **P Value** |
| **IgM – S1** | Maternal Ab concentrations between trimesters from infections during the 2nd wave | Kruskall–Wallis one-way ANOVA | P = 0.02<br>P = 0.04 |
| | Fetal Ab concentrations between trimesters from infections during the 2nd wave | Kruskall–Wallis one-way ANOVA | P ≤ 0.008 |

| | | | |
|---|---|---|---|
| | Maternal Ab concentrations between trimesters from infections during the 3rd wave | Wilcoxon Rank Sum Test | P = 0.06 |
| | Fetal Ab concentrations between trimesters from infections during the 3rd wave | Wilcoxon Rank Sum Test | P = 0.26 |
| | Maternal Ab concentrations between waves from 2nd trimester infections | Wilcoxon Rank Sum Test | P = 0.05 |
| | Fetal Ab concentrations between waves from 2nd trimester infections | Wilcoxon Rank Sum Test | P = 0.17 |
| | Maternal Ab concentrations between waves from 3rd trimester infections | Wilcoxon Rank Sum Test | P = 0.01 |
| | Fetal Ab concentrations between waves from 3rd trimester infections | Wilcoxon Rank Sum Test | P = 0.001 |
| **IgM – S2** | Maternal Ab concentrations between trimesters from infections during the 2nd wave | Kruskall–Wallis one-way ANOVA | P = 0.08 |
| | Fetal Ab concentrations between trimesters from infections during the 2nd wave | Kruskall–Wallis one-way ANOVA | P = 0.09 |
| | Maternal Ab concentrations between trimesters from infections during the 3rd wave | Wilcoxon Rank Sum Test | P = 0.003 |
| | Fetal Ab concentrations between trimesters from infections during the 3rd wave | Wilcoxon Rank Sum Test | P = 0.46 |
| | Maternal Ab concentrations between waves from 2nd trimester infections | Wilcoxon Rank Sum Test | P = 0.002 |
| | Fetal Ab concentrations between waves from 2nd trimester infections | Wilcoxon Rank Sum Test | P = 0.49 |
| | Maternal Ab concentrations between waves from 3rd trimester infections | Wilcoxon Rank Sum Test | P = 0.19 |
| | Fetal Ab concentrations between waves from 3rd trimester infections | Wilcoxon Rank Sum Test | P = 0.02 |
| **IgM –RBD** | Maternal Ab concentrations between trimesters from infections during the 2nd wave | Kruskall–Wallis one-way ANOVA | P = 0.100 |
| | Fetal Ab concentrations between trimesters from infections during the 2nd wave | Kruskall–Wallis one-way ANOVA | P = 0.17 |
| | Maternal Ab concentrations between trimesters from infections during the 3rd wave | Wilcoxon Rank Sum Test | P = 0.14 |
| | Fetal Ab concentrations between trimesters from infections during the 3rd wave | Wilcoxon Rank Sum Test | P = 0.49 |
| | Maternal Ab concentrations between waves from 2nd trimester infections | Wilcoxon Rank Sum Test | P = 0.07 |

| | | | |
|---|---|---|---|
| | Fetal Ab concentrations between waves from 2nd trimester infections | Wilcoxon Rank Sum Test | P = 0.46 |
| | Maternal Ab concentrations between waves from 3rd trimester infections | Wilcoxon Rank Sum Test | P = 0.001 |
| | Fetal Ab concentrations between waves from 3rd trimester infections | Wilcoxon Rank Sum Test | P = 0.08 |
| **IgM – N** | Maternal Ab concentrations between trimesters from infections during the 2nd wave | Kruskall–Wallis one-way ANOVA | P = 0.23 |
| | Fetal Ab concentrations between trimesters from infections during the 2nd wave | Kruskall–Wallis one-way ANOVA | P = 0.71 |
| | Maternal Ab concentrations between trimesters from infections during the 3rd wave | Wilcoxon Rank Sum Test | P = 0.01 |
| | Fetal Ab concentrations between trimesters from infections during the 3rd wave | Wilcoxon Rank Sum Test | P = 0.51 |
| | Maternal Ab concentrations between waves from 2nd trimester infections | Wilcoxon Rank Sum Test | P = 0.001 |
| | Fetal Ab concentrations between waves from 2nd trimester infections | Wilcoxon Rank Sum Test | P = 0.73 |
| | Maternal Ab concentrations between waves from 3rd trimester infections | Wilcoxon Rank Sum Test | P = 0.61 |
| | Fetal Ab concentrations between waves from 3rd trimester infections | Wilcoxon Rank Sum Test | P = 0.85 |

**Supplementary Table 2.** General AOV/AOCV procedure was used to evaluate the effect of the wave, trimester, and wave x trimester interactions on IgG and IgM Ab concentrations (S1, S2, RBD, N) in maternal and cord blood samples. P values are given for each factor and Ab.

| Ab | Wave | Trimester | Wave x Trimester interaction |
|---|---|---|---|
| **Maternal IgG** | | | |
| **S1** | 0.001 | <0.0001 | 0.001 |
| **S2** | <0.0001 | <0.0001 | <0.0001 |
| **RBD** | 0.001 | <0.0001 | 0.02 |
| **N** | 0.0004 | <0.0001 | 0.001 |
| **Maternal IgM** | | | |
| **S1** | 0.002 | 0.007 | 0.28 |
| **S2** | 0.002 | 0.005 | 0.03 |
| **RBD** | 0.002 | 0.07 | 0.43 |
| **N** | 0.13 | 0.05 | 0.02 |
| **Fetal IgG** | | | |
| **S1** | 0.17 | 0.16 | 0.10 |
| **S2** | 0.19 | 0.35 | 0.17 |
| **RBD** | 0.19 | 0.10 | 0.16 |
| **N** | 0.36 | 0.10 | 0.15 |
| **Fetal IgM** | | | |
| **S1** | 0.09 | 0.19 | 0.54 |
| **S2** | 0.29 | 0.50 | 0.40 |
| **RBD** | 0.23 | 0.28 | 0.35 |
| **N** | 0.75 | 0.40 | 0.70 |

**Supplementary Table 3.** Statistical analysis of maternal and fetal serological response to vaccination across gestation. Analysis of the data presented in Supplementary Figure 2.

| Ab Type | Comparison | Statistical Test | P Value |
|---|---|---|---|
| **IgG – S1** | Maternal Ab concentrations between trimesters of first dose vaccination | Wilcoxon Rank Sum Test | P = 0.001 |
| | Fetal Ab concentrations between trimesters of first dose vaccination | Wilcoxon Rank Sum Test | P = 0.39 |
| **IgG – S2** | Maternal Ab concentrations between trimesters of first dose vaccination | Wilcoxon Rank Sum Test | P = 0.12 |
| | Fetal Ab concentrations between trimesters of first dose vaccination | Wilcoxon Rank Sum Test | P = 0.05 |
| **IgG – RBD** | Maternal Ab concentrations between trimesters of first dose vaccination | Wilcoxon Rank Sum Test | P = 0.0001 |
| | Fetal Ab concentrations between trimesters of first dose vaccination | Wilcoxon Rank Sum Test | P = 0.97 |
| **IgG – N** | Maternal Ab concentrations between trimesters of first dose vaccination | Wilcoxon Rank Sum Test | P = 0.0001 |
| | Fetal Ab concentrations between trimesters of first dose vaccination | Wilcoxon Rank Sum Test | P < 0.0001 |
| **Ab Type** | **Comparison** | **Statistical Test** | **P Value** |
| **IgM – S1** | Maternal Ab concentrations between trimesters of first dose vaccination | Wilcoxon Rank Sum Test | P = 0.004 |
| | Fetal Ab concentrations between trimesters of first dose vaccination | Wilcoxon Rank Sum Test | P < 0.0001 |
| **IgM – S2** | Maternal Ab concentrations between trimesters of first dose vaccination | Wilcoxon Rank Sum Test | P = 0.41 |
| | Fetal Ab concentrations between trimesters of first dose vaccination | Wilcoxon Rank Sum Test | P < 0.0001 |
| **IgM – RBD** | Maternal Ab concentrations between trimesters of first dose vaccination | Wilcoxon Rank Sum Test | P = 0.61 |
| | Fetal Ab concentrations between trimesters of first dose vaccination | Wilcoxon Rank Sum Test | P = 0.01 |
| **IgM – N** | Maternal Ab concentrations between trimesters of first dose vaccination | Wilcoxon Rank Sum Test | P = 0.11 |
| | Fetal Ab concentrations between trimesters of first dose vaccination | Wilcoxon Rank Sum Test | P = 0.0004 |

**Supplementary Table 4.** Statistical analysis of maternal to fetal IgG transfer ratios across gestation. Analysis of the data presented in Supplementary Figure 3.

| Ab Type | Comparison | Statistical Test | P Value |
|---|---|---|---|
| **IgG – HA (Phuket)** | Transfer ratio between trimesters from infections during the 2nd wave | Kruskall–Wallis one-way ANOVA | P = 0.56 |
| | Transfer ratio between trimesters from infections during the 3rd wave | Wilcoxon Rank Sum Test | P = 0.67 |
| | Transfer ratio between the trimester of vaccination | Wilcoxon Rank Sum Test | P = 0.45 |
| | Transfer ratio from 2nd trimester between groups | Kruskall–Wallis one-way ANOVA, Dunn's All-Pairwise | P = 0.76 |
| | Transfer ratio from 3rd trimester between groups | Kruskall–Wallis one-way ANOVA, Dunn's All-Pairwise | P = 0.34 |
| **IgG – HA (Singapore)** | Transfer ratio between trimesters from infections during the 2nd wave | Kruskall–Wallis one-way ANOVA | P = 0.53 |
| | Transfer ratio between trimesters from infections during the 3rd wave | Wilcoxon Rank Sum Test | P = 0.98 |
| | Transfer ratio between the trimester of vaccination | Wilcoxon Rank Sum Test | P = 0.73 |
| | Transfer ratio from 2nd trimester between groups | Kruskall–Wallis one-way ANOVA, Dunn's All-Pairwise | P = 0.79 |
| | Transfer ratio from 3rd trimester between groups | Kruskall–Wallis one-way ANOVA, Dunn's All-Pairwise | P = 0.95 |
| **IgG – S1** | Transfer ratio between trimesters from infections during the 2nd wave | Kruskall–Wallis one-way ANOVA | P = 0.20 |
| | Transfer ratio between trimesters from infections during the 3rd wave | Wilcoxon Rank Sum Test | P = 0.45 |
| | Transfer ratio between the trimester of vaccination | Wilcoxon Rank Sum Test | P < 0.0001 |
| | Transfer ratio from 2nd trimester between groups | Kruskall–Wallis one-way ANOVA, Dunn's All-Pairwise | P = 0.55 |
| | Transfer ratio from 3rd trimester between groups | Kruskall–Wallis one-way ANOVA, Dunn's All-Pairwise | P = 0.16 |
| **IgG – S2** | Transfer ratio between trimesters from infections during the 2nd wave | Kruskall–Wallis one-way ANOVA | P = 0.49 |
| | Transfer ratio between trimesters from infections during the 3rd wave | Wilcoxon Rank Sum Test | P = 0.89 |
| | Transfer ratio between the trimester of vaccination | Wilcoxon Rank Sum Test | P < 0.0001 |

| | | | |
|---|---|---|---|
| | Transfer ratio from $2^{nd}$ trimester between groups | Kruskall–Wallis one-way ANOVA, Dunn's All-Pairwise | P = 0.02 |
| | Transfer ratio from $3^{rd}$ trimester between groups | Kruskall–Wallis one-way ANOVA, Dunn's All-Pairwise | P = 0.33 |
| **IgG – RBD** | Transfer ratio between trimesters from infections during the $2^{nd}$ wave | Kruskall–Wallis one-way ANOVA | P = 0.10 |
| | Transfer ratio between trimesters from infections during the $3^{rd}$ wave | Wilcoxon Rank Sum Test | P = 0.99 |
| | Transfer ratio between the trimester of vaccination | Wilcoxon Rank Sum Test | P < 0.0001 |
| | Transfer ratio from $2^{nd}$ trimester between groups | Kruskall–Wallis one-way ANOVA, Dunn's All-Pairwise | P = 0.19 |
| | Transfer ratio from $3^{rd}$ trimester between groups | Kruskall–Wallis one-way ANOVA, Dunn's All-Pairwise | P < 0.0001 |
| **IgG – N** | Transfer ratio between trimesters from infections during the $2^{nd}$ wave | Kruskall–Wallis one-way ANOVA | P = 0.006 |
| | Transfer ratio between trimesters from infections during the $3^{rd}$ wave | Wilcoxon Rank Sum Test | P = 0.80 |
| | Transfer ratio between the trimester of vaccination | Wilcoxon Rank Sum Test | P = 0.32 |
| | Transfer ratio from $2^{nd}$ trimester between groups | Kruskall–Wallis one-way ANOVA, Dunn's All-Pairwise | P = 0.03 |
| | Transfer ratio from $3^{rd}$ trimester between groups | Kruskall–Wallis one-way ANOVA, Dunn's All-Pairwise | P = 0.31 |

**Supplementary Table 5.** Values of variance and standard deviation (SD) are provided for each ab and each group. First and second row within each cell are the variance and SD values, respectively. [a,b,c] Within a row, values without a common superscript differed ($P < 0.05$).

| Ab | Control | PCR-Positive 2nd Wave | PCR-Positive 3rd Wave | Vaccinated |
|---|---|---|---|---|
| Maternal IgG – HA(Phuket) | 0.0246[a]<br>0.1568 | 0.0270[a]<br>0.1644 | 0.0278[a]<br>0.1667 | 0.0393[a]<br>0.1981 |
| Fetal IgG – HA(Phuket) | 0.0163[a]<br>0.1275 | 0.3250[b,c]<br>0.5701 | 0.4152[b]<br>0.6444 | 0.0255[a,c]<br>0.1598 |
| Maternal IgG – HA(Singapore) | 0.0842[a]<br>0.2902 | 0.0600[a]<br>0.2449 | 0.0667[a]<br>0.2582 | 0.0716[a]<br>0.2676 |
| Fetal IgG – HA(Singapore) | 0.0667[a]<br>0.2583 | 0.3029[b]<br>0.5504 | 0.3453[b]<br>0.5876 | 0.0362[a,c]<br>0.1901 |
| Maternal IgG – S1 | 0.9559[a]<br>0.9777 | 0.4491[b]<br>0.6702 | 0.5369[b]<br>0.7327 | 0.5299[b]<br>0.7280 |
| Fetal IgG – S1 | 1.1282[a]<br>1.0622 | 0.5646[b]<br>0.7514 | 0.7103[a,b]<br>0.8428 | 0.5936[b]<br>0.7705 |
| Maternal IgG – S2 | 0.4960[a]<br>0.7043 | 0.1229[b]<br>0.3505 | 0.2117[b]<br>0.4602 | 0.1626[b]<br>0.4033 |
| Fetal IgG – S2 | 0.5747[a]<br>0.7581 | 0.2891[b]<br>0.5377 | 0.4869[a,b]<br>0.6978 | 0.1562[b]<br>0.3952 |
| Maternal IgG - RBD | 0.5281[a]<br>0.7267 | 0.2420[b]<br>0.4919 | 0.2984[b]<br>0.5462 | 0.2570[b]<br>0.5069 |
| Fetal IgG - RBD | 0.7016[a]<br>0.8376 | 0.3769[b]<br>0.6140 | 0.4436[a,b]<br>0.6660 | 0.2694[b]<br>0.5190 |
| Maternal IgG - N | 0.3957[a]<br>0.6291 | 0.3020[a]<br>0.5495 | 0.3484[a]<br>0.5903 | 0.3271[a]<br>0.5719 |
| Fetal IgG - N | 0.4882[a]<br>0.6987 | 0.3389[a]<br>0.5821 | 0.3218[a]<br>0.5672 | 0.4050[a]<br>0.6364 |
